# Do dietary and physical activity interventions in adolescents provide a cost-effective use of healthcare resources? Model development and illustration based on the Engaging Adolescents in Changing Behaviour (EACH-B) programme

**DOI:** 10.1101/2021.05.05.21256672

**Authors:** Neelam Kalita, Keith Cooper, Janis Baird, Kathryn Woods-Townsend, Keith M Godfrey, Cyrus Cooper, Hazel Inskip, Mary Barker, Joanne Lord, the EACH-B study group

## Abstract

**Objective:** To assess costs, health outcomes and cost-effectiveness of interventions that aim to improve quality of diet and level of physical activity in adolescents.

**Design:** A Markov model was developed to assess four potential benefits of healthy behaviour for adolescents: better mental health, Type 2 diabetes, higher earnings and reduced incidences of adverse pregnancy outcomes. The model parameters were informed by published literature. The analysis took a societal perspective over a 20-year period. One-way and probabilistic sensitivity analyses were conducted.

**Setting:** Secondary schools.

**Participants:** A hypothetical cohort of adolescents aged 12-13 years.

**Interventions:** An exemplar school-based, multi-component intervention that was developed by the Engaging Adolescents for Changing Behaviour programme, compared with usual schooling.

**Primary and secondary outcome measures:** Incremental cost-effectiveness ratio as measured by cost per quality-adjusted life year (QALY) gained.

**Results:** The model suggested that an intervention for improving diet and physical activity has the potential to offer a cost-effective use of healthcare resources for adolescents in the UK at a willingness-to-pay threshold of £20,000 per QALY. The key model drivers are the intervention effect on levels of physical activity, quality of life gain for high levels of physical activity, the duration of the intervention effects and the period over which effects wane.

**Conclusions:** The model focused on short to medium-term benefits of healthy eating and physical activity exploiting the strong evidence base that exists for this age group. Other benefits in later life, such as reduced cardiovascular risk, are more sensitive to assumptions about the persistence of behavioural change and discounting.

**ARTICLE SUMMARY:** *Strengths and limitations of this study:*

- The study addresses an important public health question by examining if interventions targeting healthy eating and doing more physical activities provide value for money from a societal perspective.
- A Markov cohort model was developed to assess the costs and benefits, expressed in terms of quality-adjusted life years, of an exemplar school-based, multi-component intervention to improve adolescents’ diet and increase their levels of physical activity.
- The study incorporates existing evidence on the effect of improvement in adolescent health behaviours on four high prevalence short-to-medium term benefits relevant to young people: improved mental health, higher earnings, improved pregnancy outcomes and prevention of Type 2 diabetes.
- Extensive sensitivity analyses were conducted to investigate the impact of uncertainty over the model assumptions and parameter inputs, thereby highlighting areas where further research would be most useful.
- A limitation of the study is that it does not estimate the long-term impacts of such interventions due to the lack of longitudinal data on lifetime trajectories of healthy diet and increased levels of physical activity.

## INTRODUCTION

Poor diet and lack of physical activity increase the risk of non-communicable diseases (NCDs), including cardiovascular diseases, Type 2 diabetes and some cancers such as breast, colon and endometrial, in part by contributing to overweight and obesity.^1 2^ Adolescence, the life stage between childhood and adulthood, is a critical period for the development of health and disease in later life.^3 4^ Compared with other age-groups, adolescents have the unhealthiest diets and most (over 80%) fail to meet national guidelines for physical activity.^5-7^ Furthermore, the proportion meeting recommended levels of physical activity has been declining, particularly among girls (from 14% to 8%, 2008-2012).^8^

The disease burden of poor diet and physical inactivity on healthcare services is significant. In the UK, poor diet and physical inactivity cost £7 billion to the National Health Service (NHS) annually.^9^ Meeting current dietary recommendations would reduce years-of-life lost to coronary heart disease by 2 million, stroke by 400,000 and Type 2 diabetes by 19,000 over 20 years.^10^

Health behaviours in adolescence track into adulthood.^5 9 11 12^ Therefore, sub-optimal diet and body composition in adolescence not only affect immediate physical and mental health but also increase the risk of NCDs in later life. Developmental plasticity in adolescence means that interventions to improve diet and levels of physical activity have the potential to reduce the trajectory of NCD risk over the life course.^13^ While many adolescents find it difficult to engage with the long-term consequences of health behaviour, motivated and engaged adolescents can improve their health behaviours.^14 15^ Evidence suggests that school-based interventions that offer combinations of peer-modelling, social support and choice, may be effective in improving diet and physical activity amongst adolescents.^16 17 18^ Furthermore there is an increasing use of digital platforms by adolescents. According to 2018 estimates, 83% of 12-15 years olds own smartphones with 99% spending an average of 20 hours per week online.^19 20^ With an explosion in the use of such platforms to influence health behaviours in young people, there are suggestions that they have potential as a complementary feature in complex interventions that aim to influence health behaviour in adolescents.^20^

Within this framework, a research programme Engaging Adolescents in CHanging Behaviour (EACH-B) was designed to develop and test an intervention to encourage UK-based school students, aged 12-13 years, to adopt healthy behaviours such as eating better and exercising more (Trial registration: ISRCTN 74109264). EACH-B involves a cluster randomised controlled trial as a test of intervention effectiveness. Further details of the trial design are given elsewhere.^20^ The research programme is funded by the National Institute for Health Research (RP-PG-0216-20004). The “LifeLab Plus” intervention developed as part of this programme is a complex three-part programme that comprises: i) an education module that teaches school students the science behind health messages through a 2-week module with a “hands-on” practical one-day visit to a teaching laboratory at University Hospital Southampton or in school while COVID-19 restrictions apply; ii) training for teachers in skills to support behaviour change; and iii) access to a specially-designed, interactive smartphone app with game features.

Despite an emerging interest in identifying and developing interventions for improving diet and physical activity levels in adolescents, there is sparse economic evidence assessing health effects and costs of such interventions. To address this gap, we developed an illustrative decision-analytic model to assess the health benefits, costs and cost-effectiveness of a multi-component intervention such as LifeLab Plus. This prototype model is designed to investigate how changes in diet quality and levels in physical activity could affect future health outcomes and costs. We used costs data from the EACH-B programme and effectiveness from published literature. The model will be updated when results from the EACH-B trial are available.

Although there is good epidemiological evidence of long-term tracking of health behaviour,^5 9 11 12^ school-based trials rarely follow up for more than a year.^1 21 22^ The persistence of intervention effects is therefore uncertain. We take a conservative approach and focus on potential impacts of improved diet and physical activity likely to manifest in the short to medium term: up to a maximum time horizon of 20 years. We also explore alternative assumptions about the persistence of effects on behaviour after trial follow-up.

## METHODS

We developed a prototype *de novo* Markov model to estimate the costs, benefits and cost-effectiveness of school-based interventions that aim to improve diet quality and levels of physical activity compared with usual schooling for a cohort of adolescents. The model focussed on four potential short to medium-term benefits of healthy eating and physical activity in this age group: better mental health outcomes, higher earnings and reduced incidences of adverse pregnancy outcomes and Type 2 diabetes.

The model assumed that improved diet quality and increased physical activity would impact these four health outcomes via reduction in BMI. Discussion with key project stakeholders reiterated these four benefits as the most relevant obesity-related effects in this population. The model did not include later life cardiovascular disease or other chronic diseases as outcomes since the likely impact on these of an intervention undertaken as an adolescent was uncertain. The model also investigated independent effects of physical activity on diabetes and depression (i.e. direct impacts not mediated by BMI). Information relating to epidemiology, mortality, effectiveness, health-related quality of life and costs was obtained from a variety of sources and used to inform the model parameters and assumptions. See Figure 1 for model illustration.

**Figure 1:**
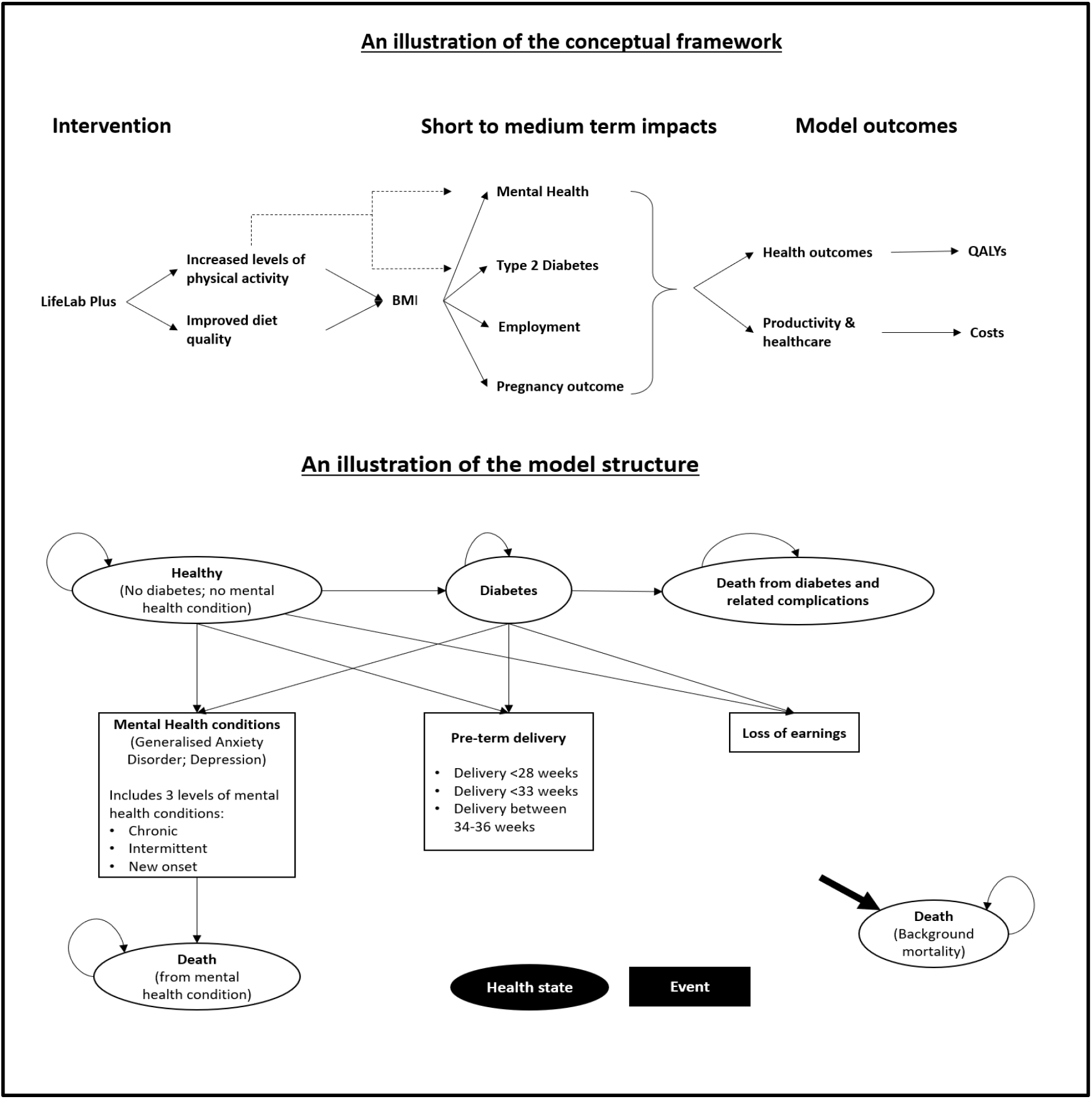
Illustration of the conceptual framework and model structure.

### Structuring the model

#### Population

A cohort of adolescents with an equal proportion of boys and girls and a mean age of 13 years was entered into the model. The intervention is based on that developed by the EACH-B programme, i.e. LifeLab Plus.^20^ The impact of this is compared with that of usual schooling.

#### Model states

The Markov model consisted of three health states: *No Type 2 Diabetes, Type 2 Diabetes* and *Death*. Outcomes associated with mental health, loss of earnings and adverse pregnancy outcome were incorporated as model events. Mental health encompasses a wide spectrum of conditions. Therefore, a pragmatic approach was adopted to include the two most relevant mental health events for adolescents: clinical depression (henceforth, referred as depression) and General Anxiety Disorder (GAD). These events were categorised as: chronic (history of persistent mental illness), intermittent (experiencing intermittent episodes), and new onset (a one-time episode). Adverse pregnancy outcome was defined by pre-term delivery categorised as: extremely pre-term (delivery <28 weeks); very pre-term (delivery <33 weeks); and moderately pre-term (delivery between 34-36 weeks). The pregnancy outcome was applicable for girls only.

Adolescents enter the model in the *No Type 2 diabetes* health state. In each model cycle, a proportion of individuals develop Type 2 diabetes and move to the *Type 2 diabetes* health state. They may also experience mental health events or pre-term births (girls only), which are treated as transient states. Each health state and event are associated with a health-related quality of life (HRQoL) and excess cost estimate. The model included intervention cost associated with LifeLab Plus and healthcare costs associated with-Type 2 diabetes, mental health events, pre-term delivery and loss of earnings due to obesity. In each model cycle, the total costs and QALYs are calculated by multiplying the individual costs and HRQoL by the number of people in the cohort still alive for each of the intervention and control arms. The total lifetime costs and QALYs are calculated by aggregating the costs and QALYs for all cycles.

#### Persistence of effects and time horizon

For the base case, the duration of the intervention effect was assumed to be sustained for 4 years with the effect waning over a period of 10 years. The model time horizon was 20 years, with yearly cycle length. This was considered appropriate as the mean age of adolescents entering the model was 13 years as it is in the EACH-B trial. The persistence of behaviour change effects from such interventions remains unclear. Therefore, the treatment effect on behaviour was not assumed to last for 20 years. Costs and outcomes were half-cycle corrected.

### Populating the model

Targeted literature searches were conducted to identify sources to inform model parameters. These are discussed below. For further details, see Appendix A and Appendix B.

#### Epidemiological data

Data on the relationship between mean BMI, age and sex was taken from Health Survey for England.^23^ For adolescents aged ≤19 years, BMI z-scores are normally used. Therefore, we rescaled the values to relate relative risks to BMI z-scores, where:

*BMI z-score = (observed value – median value of the reference population) / standard deviation value of reference population*.

Physical activity levels for children aged 13-15 years were taken from Health Survey England, 2015.^24^ The incidence of Type 2 diabetes in UK was based on an analysis of longitudinal electronic health records in the Health Improvement Network (THIN) primary care database.^25^ The prevalence of depressive episodes and GAD was taken from the Adult Psychiatric Morbidity Survey 2014^26^ and from Mental Health of Children and Young people in England 2017.^27^ The proportion in each category was assumed as follows: 17% had a chronic and 40% had a fluctuating (intermittent) course, while 43% remitted (new one-time episode).^28^ Those individuals with depression or anxiety are at higher risk of suicide than the general population.^29^ The excess death rate for those with depression and anxiety was calculated by using the suicide rate in the UK from Office of National Statistics (ONS) 2017 and applying a relative risk of 10.9 for depression and anxiety.^29^ The proportions of pre-term deliveries, obtained from ONS 2017 data, were assumed as follows: 0.5% of total births as extremely pre-term (<28 weeks), 1.2% very pre-term birth (28 to <33 weeks) and 6.3% moderately pre-term birth (33-36 weeks) respectively.^30^

#### Relationship between BMI and risks of health events

The economic model assumed a positive correlation between increased BMI and the risks of - Type 2 diabetes, depression and GAD, pre-term delivery and loss of earnings. We fitted equations to the BMI relative risks. Hazard ratios, obtained from the Medicare Current Beneficiary Survey 1991-2010, were used to estimate the increased risk of individuals with higher BMI developing Type 2 diabetes.^31^ The odds of depression and GAD in obese and overweight adolescents compared with normal-weight adolescents were obtained from Sutaria et al.^32^ This systematic review included 22 observational studies published between 2000 and 2017, representing 143,603 children. The relative risk of pre-term birth for mothers with overweight and obesity was obtained from Mcdonald et al.^33^ The risk was assumed to be the same for all three type of pre-term births.

#### Relationship between physical activity and risk of health event

The direct effect of physical activity on developing depression is modelled independent of the effect of physical activity via BMI. The odds ratio of developing depression was assumed to be 0.83 (95% CI 0.79-0.88) in those with high levels of physical activity compared to those with lower levels.^34^ Furthermore, an increase from being inactive to achieving the recommended physical activity level (150 minutes of moderate-intensity aerobic activity per week) was assumed to lower the risk of Type 2 diabetes incidence by 26%, after adjustment for body weight.^35^ The pooled odds ratio between Type 2 diabetes and risk of depression was 1.33 (95% CI, 1.18-1.51).^36^

#### Intervention effect

The intervention effect was based on three systematic reviews and meta-analyses that estimated the overall effects of school-based obesity prevention interventions.^37 38 39^ The results of the meta-analyses were found to be significantly different between groups based on BMI (− 0.17 (95% CI− 0.29, − 0.06) kg/m^2^)^38^ and BMI z-score (− 0.06 (95% CI -0.10, − 0.03))^39^ for multi-component interventions including physical activity, health education and dietary improvement. The effect of school-based intervention was assumed to increase the level of moderate or vigorous physical activity in children and adolescents by 4.84 min/day (95% CI −0.94 to 10.61).^40^

#### Mortality

General population mortality, adjusted for age and gender, was based on ONS 2020.^41^ A hazard ratio of mortality of 2.98 was applied for individuals with Type 2 diabetes and aged <65 years.^42^

#### Quality of life

EQ-5D estimates adjusted for age, gender and BMI were used to estimate quality of life. See Table 1. These estimates were obtained from 14,117 participants aged ≥16 years from the Health Survey for England 2008.^43^ Adults with diabetes was assumed to have a disutility of -0.161 where the pooled mean EQ-5D score for individuals with Type 2 diabetes was 0.67 at a mean age of 60 years.^44^ We estimated the disutility by comparing the mean general population EQ-5D score with that for diabetes. A disutility of -0.087 is used for adolescents with Type 2 diabetes based on a Swedish cohort of adolescents aged 13-18 years.^45^ A utility decrement of 0.188 was assumed for those with intermittent episodes of mental health conditions;^46^ a decrement of 0.488 for those with persistent/chronic depression;^47^ a decrement of 0.094 (half of the decrement for intermittent depression) was assumed for those with a new episode of depression. For pre-term delivery, a mean utility decrement of 0.066 was applied throughout the model time horizon. This was based on a systematic review and meta-analysis for health utility values associated with pre-term birth where all but one study used Health Utilities Index (HUI) Mark 2 (HUI2) or Mark 3 (HUI3) measures as their primary health utility assessment method.^48^ We found no evidence for quality of life loss in parents of pre-term babies. Therefore, we assumed that the quality of life decrement would be similar to intermittent mental health condition and lasts for the first two years.

**Table 1.**
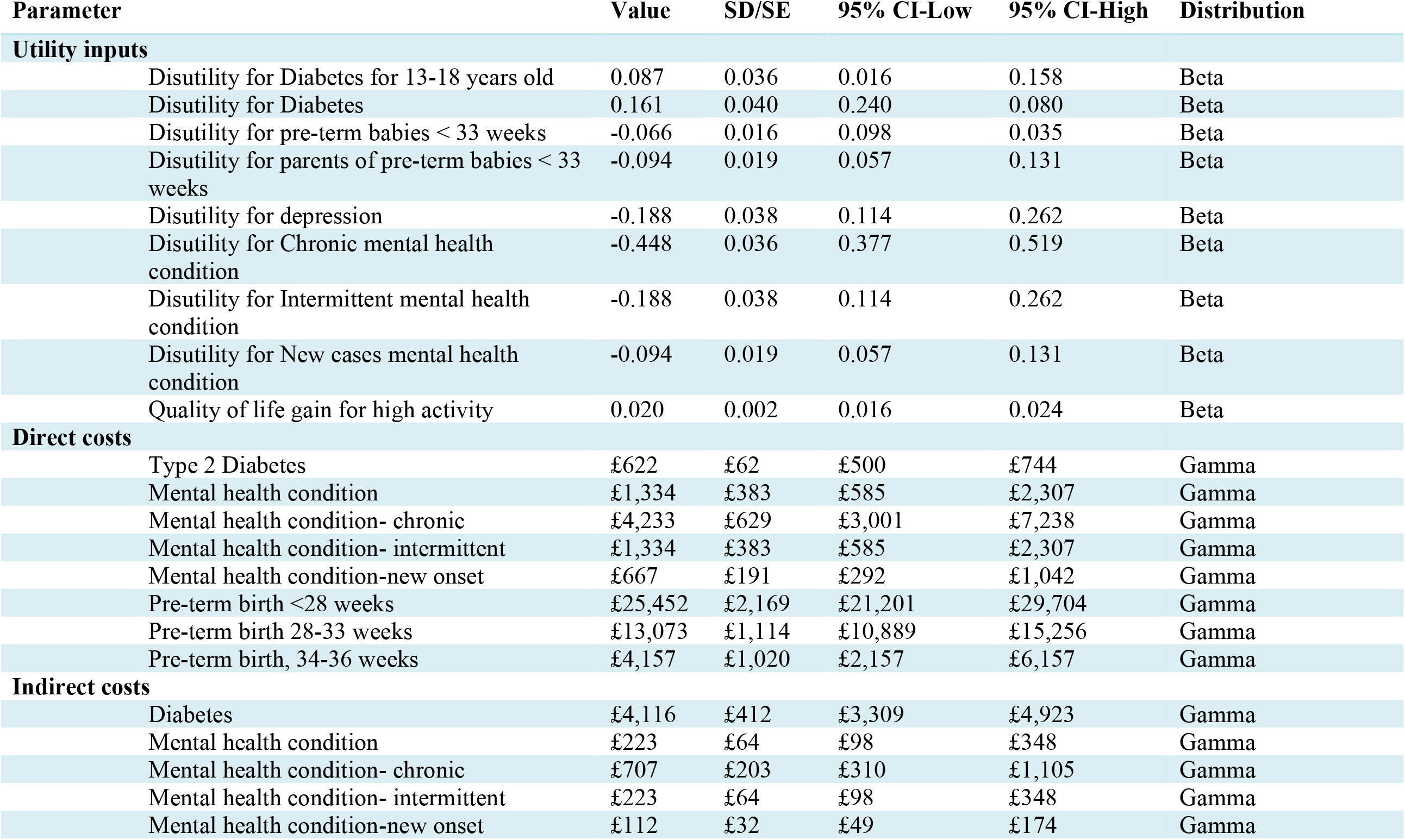
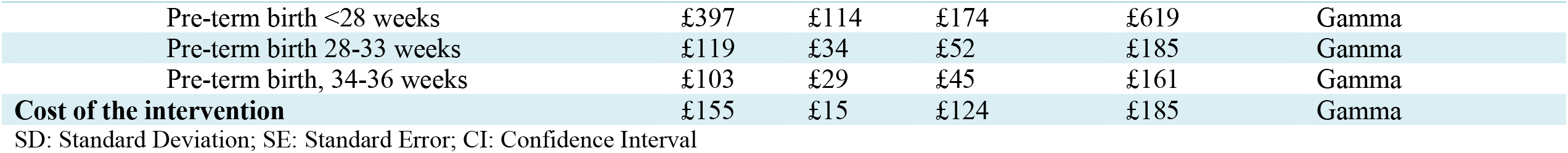
Input parameters used in the model.

| Parameter | Value | SD/SE | 95% CI-Low | 95% CI-High | Distribution |
| --- | --- | --- | --- | --- | --- |
| **Utility inputs** |  |  |  |  |  |
| Disutility for Diabetes for 13-18 years old | 0.087 | 0.036 | 0.016 | 0.158 | Beta |
| Disutility for Diabetes | 0.161 | 0.040 | 0.240 | 0.080 | Beta |
| Disutility for pre-term babies < 33 weeks | -0.066 | 0.016 | 0.098 | 0.035 | Beta |
| Disutility for parents of pre-term babies < 33 weeks | -0.094 | 0.019 | 0.057 | 0.131 | Beta |
| Disutility for depression | -0.188 | 0.038 | 0.114 | 0.262 | Beta |
| Disutility for Chronic mental health condition | -0.448 | 0.036 | 0.377 | 0.519 | Beta |
| Disutility for Intermittent mental health condition | -0.188 | 0.038 | 0.114 | 0.262 | Beta |
| Disutility for New cases mental health condition | -0.094 | 0.019 | 0.057 | 0.131 | Beta |
| Quality of life gain for high activity | 0.020 | 0.002 | 0.016 | 0.024 | Beta |
| **Direct costs** |  |  |  |  |  |
| Type 2 Diabetes | £622 | £62 | £500 | £744 | Gamma |
| Mental health condition | £1,334 | £383 | £585 | £2,307 | Gamma |
| Mental health condition- chronic | £4,233 | £629 | £3,001 | £7,238 | Gamma |
| Mental health condition- intermittent | £1,334 | £383 | £585 | £2,307 | Gamma |
| Mental health condition-new onset | £667 | £191 | £292 | £1,042 | Gamma |
| Pre-term birth <28 weeks | £25,452 | £2,169 | £21,201 | £29,704 | Gamma |
| Pre-term birth 28-33 weeks | £13,073 | £1,114 | £10,889 | £15,256 | Gamma |
| Pre-term birth, 34-36 weeks | £4,157 | £1,020 | £2,157 | £6,157 | Gamma |
| **Indirect costs** |  |  |  |  |  |
| Diabetes | £4,116 | £412 | £3,309 | £4,923 | Gamma |
| Mental health condition | £223 | £64 | £98 | £348 | Gamma |
| Mental health condition- chronic | £707 | £203 | £310 | £1,105 | Gamma |
| Mental health condition- intermittent | £223 | £64 | £98 | £348 | Gamma |
| Mental health condition-new onset | £112 | £32 | £49 | £174 | Gamma |

**Table 1 Input parameters used in the model**
| **Parameter** | **Value** | **SD/SE** | **95% CI-Low** | **95% CI-High** | **Distribution** |
| --- | --- | --- | --- | --- | --- |
| Pre-term birth <28 weeks | £397 | £114 | £174 | £619 | Gamma |
| Pre-term birth 28-33 weeks | £119 | £34 | £52 | £185 | Gamma |
| Pre-term birth, 34-36 weeks | £103 | £29 | £45 | £161 | Gamma |
| **Cost of the intervention** | £155 | £15 | £124 | £185 | Gamma |

SD: Standard Deviation; SE: Standard Error; CI: Confidence Interval

#### Costs

##### Intervention costs

The intervention cost for our illustrative analysis was based on LifeLab Plus. Further information on resources used for delivering LifeLab are in Appendix B. The cost of the application used delivered as part of the intervention, was incorporated as a capital cost and was assumed to last 10 years and be used in 10 centres. Similarly, the cost of setting up LifeLab Plus in a different centre was assumed to consist of one year’s staff costs and assumed to last for 10 years. Maintenance costs were estimated at 25% of the development cost per year. Overheads were included according to the rates used in Curtis et al. 2019^49^, i.e. direct overheads based on 29% of direct care salary costs and indirect overheads based on 16% direct care salary costs.

##### Health state costs

Literature for costing studies and national datasets relating to individuals with Type 2 diabetes, depression, pre-term birth, and loss of earnings related to obesity were reviewed to inform the cost parameters. Both direct and indirect costs were included. Indirect costs included the effect of depression on income and productivity. Costs were updated to 2019 prices using the hospital and community health services index.^49^ See Table 1.

The current and future costs for Type 2 diabetes were sourced from Hex et al.^50^ Direct health costs and indirect societal and productivity costs were estimated using a top-down approach. We assumed that individuals with diabetes would not incur costs for complications as these are likely to affect individuals who have diabetes for a longer period. Direct and indirect costs for depression were taken from a trial of patients with a history of recurrent depression by Kuyken et al.;^51^ and the costs associated with pre-term birth were based on the study by Khan et al.^52^

Total societal costs for children born at 32-33 weeks and 34-36 weeks gestation from birth to 24 months were taken from a study that compared these costs to those for children born at full-term. Costs for children born <28 weeks and 28-33 weeks were estimated, assuming that they varied in the same way.^53^ The cost used in the model was the mean societal cost difference between these two groups.

The lifetime indirect costs for overweight and obesity in childhood and adolescence is based on the study by Hamilton et al.^54^ Mean total lifetime healthcare and productivity costs were estimated in Irish Euros, which were converted to GBP (£) and adjusted according to the average wage in the UK.

#### Validation

The structure of the prototype model was validated by the study team comprising epidemiologists, statisticians, trialists, public health experts and health economists. Internal validity of the model was established through running several tests and ensuring that the model predictions were consistent with the model specification.

#### Cost-effectiveness analysis

We followed the modelling guidelines advocated by NICE Reference case,^55^ with an exception for the perspective adopted for the analysis. The economic evaluation was conducted from a societal perspective that included both direct and indirect costs. Costs and benefits were discounted at 3.5% per year and expressed in terms of QALYs. These were estimated and averaged across the simulated cohort. The incremental cost-effectiveness ratio (ICER) was estimated as a ratio of the incremental costs of the intervention, i.e. LifeLab Plus relative to the comparator (usual schooling) to the incremental QALYs of the intervention relative to the comparator. The intervention was considered cost-effective if the ICER was below the lower (more conservative) threshold of £20,000 per QALY gained, as recommended for the English NHS by NICE.^55^

#### Sensitivity analyses

To assess the uncertainty around the model predictions, we conducted deterministic sensitivity analysis (DSA), probabilistic sensitivity analyses (PSA) and scenario analyses. For the DSA, input parameters were varied by 95% confidence intervals where available. Monte Carlo simulations of 1000 iterations were run for the PSA to assess the combined effects of input parameter uncertainties where parameters were simultaneously sampled within a specified distribution. See Table 1. Further details in Appendix A. Scenario analyses were conducted to assess structural uncertainties related to model assumptions. Uncertainty about the sustainability of the intervention effect was assessed by varying the duration of intervention effect and its waning period.

Effectiveness parameters were assigned beta and lognormal distributions, utilities were assumed to follow a beta distribution, and costs were assigned a gamma distribution. The model was developed and implemented in Microsoft Excel.

#### Patient and public involvement

The research questions addressed in the overarching research programme EACH-B were informed by public involvement. Furthermore, representatives from public involved in the EACH-B research programme were presented the conceptual framework, modelling approaches and invited to comment.

## RESULTS

### Base case analysis

In the base case analysis, the exemplar intervention based on the multi-component LifeLab Plus was associated with higher costs and better health outcomes (more QALYs) compared with usual schooling. LifeLab Plus was associated with a mean QALY gain of 0.0085 at an incremental cost of £123 per person compared with usual schooling, resulting in an ICER of £14,367 per QALY. See Table 2.

**Table 2.** Incremental cost-effectiveness ratios in the base case and scenario analyses.

|  | **Total cost (£)** | **Total QALYs** | **Incremental cost (£)** | **Incremental QALY** | **ICER** |
| --- | --- | --- | --- | --- | --- |
| **Base case analysis** |  |  |  |  |  |
| Usual schooling | £4,416 | 13.07 |  |  |  |
| LifeLab Plus | £4,539 | 13.08 | £123 | 0.0085 | £14,367 |
| **Perspective: NHS** |  |  |  |  |  |
| Usual schooling | £2,001 | 13.07 |  |  |  |
| LifeLab Plus | £2,141 | 13.08 | £140 | 0.0085 | £16,339 |
| **Costs discounted at 3.5%; QALYs at 1.5% per annum** |  |  |  |  |  |
| Usual schooling | £4,416 | 15.76 |  |  |  |
| LifeLab Plus | £4,539 | 15.77 | £123 | 0.0097 | £12,640 |
| **Costs and QALYs discounted at 1.5%** |  |  |  |  |  |
| Usual schooling | £5,813 | 15.76 |  |  |  |
| LifeLab Plus | £5,930 | 15.77 | £117 | 0.0097 | £12,044 |
| **Time Horizon: 5 years** |  |  |  |  |  |
| Usual schooling | £488 | 4.25 |  |  |  |
| LifeLab Plus | £633 | 4.25 | £145 | 0.0043 | £33,423 |
| **Time Horizon: 10 years** |  |  |  |  |  |
| Usual schooling | £1,132 | 7.76 |  |  |  |
| LifeLab Plus | £1,265 | 7.77 | £134 | 0.0071 | £18,851 |
| **Time Horizon: 30 years** |  |  |  |  |  |
| Usual schooling | £11,137 | 16.67 |  |  |  |
| LifeLab Plus | £11,258 | 16.67 | £121 | 0.0088 | £13,719 |
| **Time Horizon: 40 years** |  |  |  |  |  |
| Usual schooling | £18,417 | 19.05 |  |  |  |
| LifeLab Plus | £18,537 | 19.06 | £120 | 0.0089 | £13,455 |
| **Duration of treatment effect: 1 year & Treatment waning: 2 years** |  |  |  |  |  |
| Usual schooling | £4,416 | 13.07 |  |  |  |
| LifeLab Plus | £4,566 | 13.08 | £150 | 0.0030 | £49,758 |
| **Duration of treatment effect: 1 years & Treatment waning: 5 years** |  |  |  |  |  |
| Usual schooling | £4,416 | 13.07 |  |  |  |
| LifeLab Plus | £4,563 | 13.08 | £147 | 0.0043 | £34,089 |
| **Duration of treatment effect: 2 years & Treatment waning: 5 years** |  |  |  |  |  |
| Usual schooling | £4,416 | 13.07 |  |  |  |
| LifeLab Plus | £4,560 | 13.08 | £144 | 0.0051 | £28,034 |
| **Duration of treatment effect: 5 years & Treatment waning: 5 years** |  |  |  |  |  |
| Usual schooling | £4,416 | 13.07 |  |  |  |
| LifeLab Plus | £4,553 | 13.08 | £137 | 0.0075 | £18,218 |
| **Duration of treatment effect: 5 years & Treatment waning: 10 years** |  |  |  |  |  |
| Usual schooling | £4,416 | 13.07 |  |  |  |
| LifeLab Plus | £4,529 | 13.08 | £113 | 0.0092 | £12,184 |
| **Duration of treatment effect: 10 years & Treatment waning: 10 years** |  |  |  |  |  |
| Usual schooling | £4,416 | 13.07 |  |  |  |
| LifeLab Plus | £4,432 | 13.09 | £16 | 0.0125 | £1,257 |
| **Duration of treatment effect: 15 years & Treatment waning: 5 years** |  |  |  |  |  |
| Usual schooling | £4,416 | 13.07 |  |  |  |

|  | **Total cost**<br>**(£)** | **Total**<br>**QALYs** | **Incremental**<br>**cost (£)** | **Incremental**<br>**QALY** | **ICER** |
| --- | --- | --- | --- | --- | --- |
| LifeLab Plus | £4,308 | 13.09 | -£108 | 0.0147 | Dominates |

ICER, incremental cost-effectiveness ratio (incremental cost/incremental QALY); QALY, quality-adjusted life year.

### Sensitivity analyses

As outlined in the methods section, sensitivity analyses were implemented by varying the base case assumptions and parameter inputs. The result of the one-way sensitivity analyses is presented as a Tornado plot. See Figure 2. Parameters such as the intervention effect on levels of physical activity (expressed in terms of minutes of moderate or vigorous physical activity), quality of life gain for high levels of physical activity, duration of intervention effect and duration of treatment waning period had the highest impact on the cost-effectiveness results. Other parameters such as effect of physical activity on BMI, time horizon and intervention costs also influenced the base case results, but to a lesser extent. The results of the PSA presented as the cost-effectiveness scatter plot showed that the simulations lie in the North-East quadrant of the cost-effectiveness plane. See Figure 3. This implies that the intervention-LifeLab Plus is likely to produce health benefits at an additional cost. At a willingness-to-pay threshold of £20,000 per QALY gained, the probability of LifeLab Plus being cost-effective was 69% compared with usual schooling at 31% respectively.

**Figure 2:**
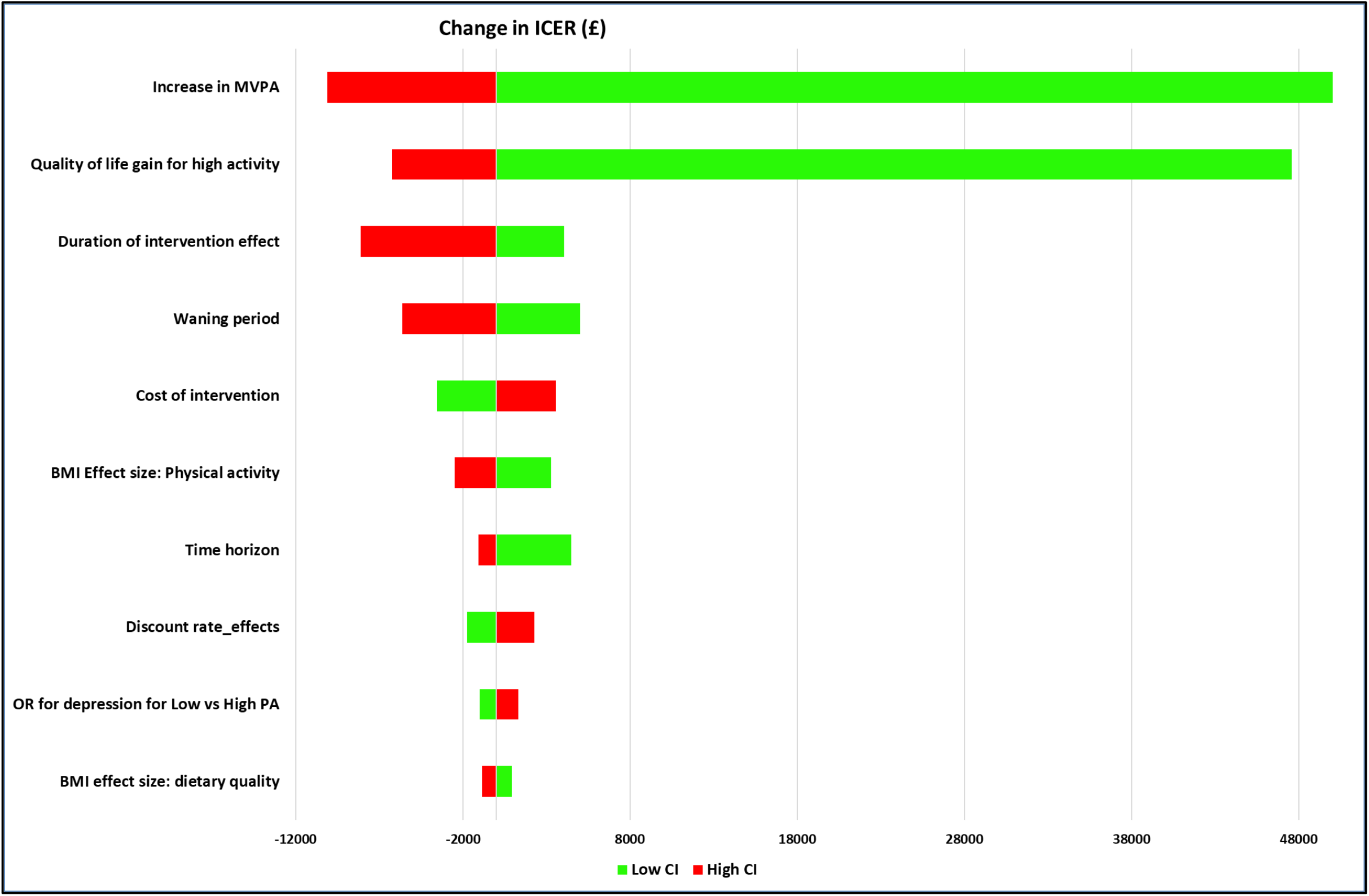
Tornado diagram to show the impact of varying parameter input values on ICERs. MVPA: Moderate or Vigorous Physical Activity; BMI: Body Mass Index; CI: Confidence Interval; ICER: Incremental Cost Effectiveness Ratio

**Figure 3:**
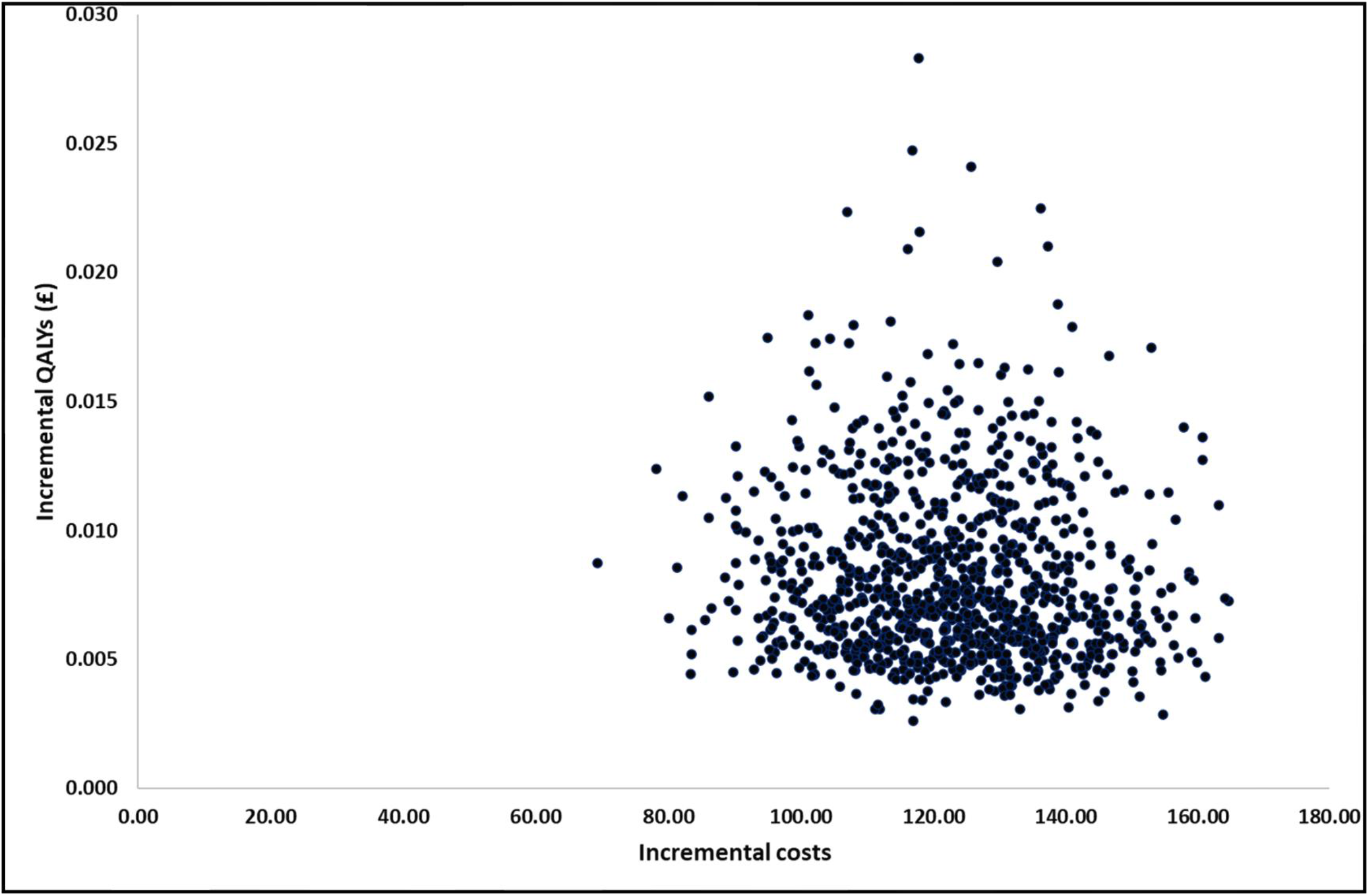
Scatter plot of PSA cost-effectiveness results.

## DISCUSSION

### Main findings

Based on economic modelling, we estimate that a multi-component intervention to improve dietary quality and physical activity, such as LifeLab Plus, would be considered cost-effective under conventional willingness-to-pay thresholds of £20,000–£30,000 per QALY gained in the UK. For our base case, the duration of the treatment effect was assumed to be sustained for 4 years with the effect waning over a further period of 10 years. Our sensitivity analyses showed that if the duration of the treatment effect was not sustained to this extent, the intervention would be less cost-effective. See Table 2.

### Comparison with previous models

Previous cost-effectiveness studies evaluating dietary and physical activity interventions for adolescents have largely consisted of within trial analyses that have not considered the benefits beyond the trial period.^56-60^ The cost-effectiveness of these intervention vary between cost saving of NZ$835 per child for the low intensity program^56^ to £120,630 per QALY for the HELP intervention.^59^ However, comparison between studies is difficult because of the differences in the study designs, the interventions considered and the outcomes reported.

Gc et al^1^ developed a model to assess the long-term costs and health outcomes of two physical activity interventions targeting adolescents in UK. The cost-effectiveness of these interventions varied between £11,426 per QALY for an after-school intervention and £68,056 per QALY for a multicomponent intervention. The costs of these interventions varied between £51 per participant for the after-school intervention to AUS$394 for the multicomponent intervention. Their model included different health states to our model for diseases which typically affect people in later life such as chronic heart disease, stroke, heart failure, breast failure and colorectal cancer. They ran the model for a lifetime horizon of 65 years. We have not included these health states as our cost-effectiveness results are based on a conservative assumption that treatment benefits for adolescents from such multicomponent intervention as LifeLab Plus do not persist beyond 20 years.

### Strengths and limitations

The study addresses an important public health question by examining if interventions targeting healthy eating and doing more physical activities provide value for money from a societal perspective. The study incorporates existing evidence on the effect of improvement in adolescent health behaviours on four high prevalence short-to-medium term benefits relevant to young people: improved mental health, higher earnings, improved pregnancy outcomes and prevention of Type 2 diabetes. Sources for data used within our model were identified from a targeted literature review. Where data were not available for adolescents, we have used data from the adult population. Model structure and assumptions were informed by this review and discussions with public health experts. UK-specific incidence rates were used to ensure that patients entering the model matched the likely distribution of events in the UK.

The model does not estimate the long-term impacts of such interventions due to the lack of longitudinal data on lifetime trajectories of healthy diet and increased levels of physical activity. If the effects are more lasting, then additional benefits such as prevention of cardiovascular disease would enhance the cost-effectiveness of the intervention. Per se our cost-effectiveness results are conservative.

Our model has several shortcomings. Firstly, there is uncertainty around key assumptions related to the duration of benefits from the intervention. In our analysis, we have assumed that the benefit observed in the clinical trials will last for 4 years and then will gradually reduce over the next 10 years. We only include costs and effects over a 20-year time horizon and have assumed that there would be no further benefits of the intervention for chronic diseases such as cardiovascular diseases and diabetes. Secondly, there are limitations in our approach to estimating income lost due to obesity. We have adopted a simple approach; however, this is a complex interacting bi-directional system. We have not fully explored whether income loss is due to obesity or whether obesity is caused by income loss through, for example, unemployment. Finally, where data from adolescent age groups were unavailable, those from adult populations were used to inform model parameters.

## CONCLUSION

Complex behavioural interventions that aim to improve diet and increase levels of physical activity amongst school-aged children have the potential to provide cost-effective use of UK healthcare resources. Such interventions have the potential to reduce burden of NCDs, although benefits in later life are more sensitive to assumptions about the persistence of behavioural change and discounting. Lastly, there is a need to establish long-term (preferably, lifetime) effectiveness of such interventions.

## Data Availability

Data from publicly available sources were used for the model. All model inputs are described in the paper.

## Author Contributions

NK designed the model, performed the analysis, interpreted the analysis, and wrote the first draft of the manuscript. KC designed the model and performed the analysis. JL supervised the process, provided comments on model structure and data analysis. KWT provided inputs for model parameters. JB, KMG, HI, MB and CC provided critical comments on the model structure. All authors contributed to the critical revision of the manuscript and approved the final version of the manuscript.

## Patient consent for publication

Not required

