## Appendix A, Appendix B, Appendix C for "Do dietary and physical activity interventions in adolescents provide a cost-effective use of healthcare resources? Model development and illustration based on the Engaging Adolescents in Changing Behaviour (EACH-B) programme"

1 Appendix A

Model parameters were estimated by using the following equations. For the control arm, rate was based upon age only. For the intervention arm, rate was based on age and the change in BMI from the control population to the intervention.

**Table 1: Parameter equations**

| **Parameter** | **Equation** | **Units** | **Source** |
| --- | --- | --- | --- |
| BMI - men | -0.0031*age^2^+0.4096*age+16.098 | Kg/m2 | HSE 2017^1^ |
| BMI - women | -0.0032*Age^2^ + 0.4235*Age+15.812 | Kg/m2 |  |
| Type 2 Diabetes Men Control, age <25y | 0.025*Age-0.265 | Incidence per 1000 | Sharma 2015^2^ |
| Type 2 Diabetes Men Control, age >25y | 0.0054Age^2-0.191Age+1.7142 | Incidence per 1000 |  |
| Type 2 Diabetes Women, Control, all ages | 0.0031Age^2-0.0942Age+1.2045 | Incidence per 1000 |  |
| Type 2 Diabetes Men, Intervention, age < 25y | (0.025*Age-0.265)/(0.081BMI_control-0.8451)*(0.081BMI_int-0.8451) | Incidence per 1000 | Gray et al 2015^3^ |
| Type 2 Diabetes Men, Intervention, > 25y | (0.0054Age^2-0.191Age+1.7142)/(0.081BMI_control-0.8451)*(0.081BMI_int-0.8451) | Incidence per 1000 |  |
| Type 2 Diabetes Women Intervention, all ages | (0.0031Age^2-0.0942Age+1.2045)/(0.0637BMI_control-0.3135)*(0.0637BMI_Int-0.3135) | Incidence per 1000 |  |
| Mental health Men, control | -0.00008*Age^2+0.0073*Age-0.0544 | % Incidence | APMS 2014^4^ |
| Mental health Women, control | -0.00007*Age^2+0.0062*Age+0.0034 | % Incidence |  |
| Mental Health Men, intervention | (-0.00007*Age^2+0.0062*Age+0.0034)*  (0.0127*BMI_I+0.727)/(0.0127*BMI_C+0.727) | % Incidence | Sutaria et al 2018^5^ |
| Mental Health Women, intervention | (-0.00007*Age^2+0.0062*Age+0.0034) *(0.0391*BMI_I+0.1087)/(0.0391*BMI_C+0.1087) | % Incidence |  |
| Mental Health, diabetes | Multiply MH risk by 1.33 |  | Chireh et al 2019^6^ |
| Mental Health, Physical exercise | OR 0.83 high activity vs low activity. Applied to intervention arm only. |  | Schuch et al 2018^7^ |
| Preterm birth <33 weeks, control | -0.0512*Age^2^+3.0922*Age-37.891 | Incidence per 1000 | ONS 2017^8^ |
| Preterm birth <33 weeks, intervention | (-0.0512*Age2+3.0922*Age-37.891)* (0.0514*BMI_I-0.1712)/ (0.0514*BMI_C-0.1712) | Incidence per 1000 | Mcdonald et al 2010^9^ |
| Loss of earnings, men | 12534BMI – 297703 | £ over lifetime | Hamilton et al 2018^10^ |
| Loss of earning, women | 17695BMI - 439796 | £ over lifetime |  |

**Table 2: Parameter equations for z-scores**

| **Parameter** | **Equation** | **Units** | **Source** |
| --- | --- | --- | --- |
| Mental Health Men, Intervention | (0.0054Age^2-0.191Age+1.7142)/  (0.0274BMI_z_score_control+1.0011)*(0.0274BMI_z_scores_int+1.011) | Incidence per 1000 | Gray et al 2015^3^ |
| Mental Health Women, Intervention | (0.0031Age^2-0.0942Age+1.2045)/(0.084BMI_z_scores_control+ 0.9486)*(0.084BMI_z_scores_Int+0.9486) | Incidence per 1000 |  |
| Type 2 Diabetes Men, intervention | (-0.00007*Age^2+0.0062*Age+0.0034)/(0.1742BMI_z_scores_control+ 0.8964)*(0.1742BMI_z_scores_Int+0.8964) | % Incidence | Sutaria et al 2018^5^ |
| Type 2 Diabetes Women, intervention | (-0.00007*Age^2+0.0062*Age+0.0034) *(0.1369*BMI_z_scores_I+1.0556)/(0.1369*BMI_z_scores_C+1.0556) | % Incidence |  |
| Preterm birth <33 weeks, intervention | (-0.0512*Age2+3.0922*Age-37.891)* (0.1105*BMI_z_scores_I+0.9336)/ (0.1105*BMI_z_scores_C+0.9336) | Incidence per 1000 | Mcdonald et al 2010^9^ |

**Table 3: Parameter equation for quality of life estimates**

| **Parameter** | **Equation** | **Source** |
| --- | --- | --- |
| Quality of life, by age | 0.9508566+0.0212126*Male-0.0002587*Age-0.0000332*Age^2^ | Maheswaran et al 2012^11^ |
| Disutility for BMI, if BMI < 30 | -0.0009BMI + 0.0189 |  |
| Disutility for BMI, if BMI > 30 | -0.0036BMI+0.094 |  |

**Table 4: Input parameters used in the model**

| Parameter | | Value | SD/SE | 95% CI-Low | 95% CI-High | Distribution |
| --- | --- | --- | --- | --- | --- | --- |
| Direct trial efficacy | |  |  |  |  |  |
|  | BMI effect size: Physical activity | 0.13 | 0.046 | 0.040 | 0.220 | Beta |
|  | BMI effect size: Dietary quality | 0.04 | 0.01423 | 0.012 | 0.068 | Beta |
|  | BMI z-scores effect size: Physical activity | 0.04 | 0.02 | 0.001 | 0.079 | Beta |
|  | BMI z-scores effect size: Dietary quality | 0.02 | 0.01 | 0.000 | 0.040 | Beta |
|  | Prop. meeting PA guidelines control | 0.13 | 0.0013 | 0.127 | 0.133 | Beta |
|  | Prop. meeting PA guidelines intervention | 0.16 | 0.049 | 0.117 | 0.257 | Beta |
|  | Increase in Moderate to Vigorous PA | 4.84 | 2.944 | -0.940 | 10.610 | Beta |
| Relative effectiveness | |  |  |  |  |  |
|  | OR for depression for Low vs High Physical Activity | 0.83 | 0.026 | 0.790 | 0.880 | Beta |
|  | OR of diabetes increasing the risk of depression | 1.33 | 0.133 | 1.069 | 1.591 | Lognormal |
|  | RR of PA on incidence of diabetes | 0.74 | 0.074 | 0.595 | 0.885 | Lognormal |
| Mortality | |  |  |  |  |  |
|  | Mental health: risk of death by suicide | 0.001 | 0.000 | 0.001 | 0.001 | Beta |
|  | HR of mortality those with type 2 Diabetes aged <65 years | 2.978 | 0.190 | 2.605 | 3.350 | Lognormal |
|  | HR of mortality those with type 2 Diabetes aged >65 years | 1.958 | 0.050 | 1.860 | 2.056 | Lognormal |
| Relative effectiveness^a^ | |  |  |  |  |  |
|  | Pre-term birth relative risk for BMI P1 | 0.051 | 0.005 | 0.041 | 0.061 | Beta |
|  | Pre-term birth relative risk for BMI P2 | -0.171 |  | 0.111 | -0.322 |  |
|  | MH BMI relative risk Boys P1 | 0.013 | 0.004 | 0.004 | 0.021 | Beta |
|  | MH BMI relative risk Boys P2 | 0.727 |  | 0.914 | 0.540 |  |
|  | MH BMI relative risk girls P1 | 0.039 | 0.004 | 0.031 | 0.047 | Beta |
|  | MH BMI relative risk girls P2 | 0.109 |  | 0.324 | -0.005 |  |
|  | Diabetes relative risk boys P1 | 0.081 | 0.004 | 0.073 | 0.089 | Beta |
|  | Diabetes relative risk boys P2 | -0.845 |  | -0.571 | -0.912 |  |
|  | Diabetes relative risk girls P1 | 0.064 | 0.013 | 0.039 | 0.089 | Beta |
|  | Diabetes relative risk girls P2 | -0.314 |  | 0.167 | -0.906 |  |
|  | Loss of earnings Men P1 | 12036 | 601.8 | 10856 | 13215.5 | Gamma |
|  | Loss of earnings Men P2 | -284478 |  | -256213 | -311886.5 |  |
|  | Loss of earnings Women P1 | 16689 | 1668.9 | 13418 | 19960.0 | Gamma |
|  | Loss of earnings Women P2 | -412424 |  | -339474 | -504989.1 |  |
| Z scores ^a^ | |  |  |  |  |  |
|  | Pre-term birth relative risk for BMI z-scores P1 | 0.1105 | 0.011 | 0.089 | 0.132 | Beta |
|  | Pre-term birth relative risk for BMI z-scores P2 | 0.9336 |  | -0.910 | -1.841 |  |
|  | MH BMI z-scores relative risk Boys P1 | 0.0274 | 0.003 | 0.022 | 0.033 | Beta |
|  | MH BMI z-scores relative risk Boys P2 | 1.0011 |  | 0.526 | 0.295 |  |
|  | MH BMI z-scores relative risk girls P1 | 0.084 | 0.008 | 0.068 | 0.100 | Beta |
|  | MH BMI z-scores relative risk girls P2 | 0.9486 |  | -0.452 | -1.160 |  |
|  | Diabetes relative risk boys P1 | 0.1742 | 0.009 | 0.157 | 0.191 | Beta |
|  | Diabetes relative risk boys P2 | 0.8964 |  | -2.378 | -3.112 |  |
|  | Diabetes relative risk girls P1 | 0.1369 | 0.027 | 0.083 | 0.191 | Beta |
|  | Diabetes relative risk girls P2 | 1.0556 |  | -0.790 | -3.097 |  |
| Proportions of population with mental health conditions | | | | | | |
|  | Chronic | 0.170 | 0.017 | 0.137 | 0.203 | Dirichlet |
|  | intermittent | 0.400 | 0.040 | 0.322 | 0.478 | Dirichlet |
|  | New and one-off | 0.430 | 0.043 | 0.346 | 0.514 | Dirichlet |
| ^a^ Risk equation is in the form: Relative risk = P1*BMI + P2 | | | | | | |

2 Appendix B

A breakdown of the resources used for estimating the costs of the intervention i.e. LifeLab Plus is presented below. The intervention comprises of three components:

i. Life Lab
ii. Digital App
iii. Teachers’ Support (Healthy Conversation skills)

The costs are estimated for one-year that is, for the trial duration. The assumptions related to resources-used are based on a discussion with KTW on 5^th^ June 2019.

**Table 4: Life Lab**

| **Item** | **Number** | **Comment** |
| --- | --- | --- |
| Number of classes per year | 50 |  |
| No of pupils in each class | 25 | Varies between 25-32; assumed 25 for the base case |
| Total pupils | 1250 | =50 classes x 25 pupils per class |
| No of schoolteachers in each class | 1 |  |
| Total teachers | 50 |  |

**Table 5: Staff Training for Life Lab and Healthy Conversation Skills**

| **Item** | **Number** | **Comment** |
| --- | --- | --- |
| Lifelab staff / training day (days) | 6 | Each training day / Lifelab day consists of 6 staff days to run. 4 staff days to run training and preparation and 2 days of administration. |
| Teacher training |  |  |
| No. of Training days | 5 | Each training day is for 10 teachers. |
| Lifelab Staff days | 30 | =5 training days x 6 staff days / training day |
| Lifelab |  |  |
| Number of days | 50 | =50x1 |
| Lifelab Staff days | 300 | =50 life lab days x 6 staff day / lifelab |
| Scientists |  | Each lifelab day consists of 3 scientists |
| Number trained | 75 | Assumed each scientist does two lifelab days |
| Training days | 3 | =75 scientists /25; 25 trained per day |
| Lifelab Staff days | 18 | =3 training days x 6 staff days / training day |
| Training the trainers (Healthy Conversation Skills) |  |  |
| Trainers | 6 | Assumption |
| Lifelab Staff days | 1 | Assume takes 2 hours |
| Showcase |  | All schools |
| Lifelab Staff days | 3.5 | 1.5 days prep; 1 day delivery for 2 staff |
| Total lifelab staff days | 352.5 | Band 4 teacher |

*Data collection – Lifelab*

Each class has two hours of data collection by two Lifelab staffs, 2 research nurses and 1 research assistant.

**Table 6: Costs for data collection**

| **Item** | **Number** | **Comment** |
| --- | --- | --- |
| Lifelab staff days | 28.6 | =2 hours x2 staff x50 (lifelab days)/7 (hours) |
| Research nurse | 28.6 | =2 hours x2 staff x50 (lifelab days)/7 (hours) |
| Research assistant | 14.3 | =2 hours x1 staff x50 (lifelab days)/7 (hours) |

**Table 7: Staff unit costs**

| **Staff costs** | **Salary** | **Comment** |
| --- | --- | --- |
| Band 4 teacher | £34,189 | i.e. £163 per day, Assumes 210 working days per year |
| Research assistant | £27,025 | i.e. £129 per day, Assumes 210 working days per year |
| Research nurse Band 5 | £26,231 | i.e £125 per day, Assumes 210 working days per year |

**Table 8: Staff total costs**

| **Staff costs** | **Total** | **Comment** |
| --- | --- | --- |
| Lifelab / training | £71,736 | =£34,189/210x352.5*1.25;  = salary / working days per year*lifelab staff days*NI and pension adjustment  Assumes 210 working days per year, pension / NI contributions of 25% |
| Follow-up data collection | £12,641 | =(£34,189*28.6 + £27,025 *28.6 + 26,231 *14.3)/210*1.25  = (salary teacher* data collection days teacher + salary research nurse* data collection days research nurse + salary research assistant*data collection days research assistant)/working days per year*NI / pension adjustment |
| Total | £84,377 |  |

**Table 9: Digital App development**

| **Item** | **Total** | **Comment** |
| --- | --- | --- |
| Development of app | £324,000 | Estimate from expert |
| Duration of development, year | 5 | As above |
| Duration of app, years | 10 | Assumption |
| Number of centres using app | 10 | Assumption |
| Cost per year development | £64,800 | £324,000 / 5; total cost / number of years of development |
| Maintenance cost | £16,200 | Assumed to be 25% of development cost |
| Cost per year of development across app lifetime per centre | £3240 | £324,000 / 100; total development cost / (number of years app lasts for x number of centres) |
| EACH-B development for new centre | £87,445 | Assumed to be 1 year of staff costs. |
| Number of years centre lasts | 10 | Assumption |
| Cost per year of centre | £8745 | £87,445 / 10 |

**Table 10: Summary of included costs**

| **Item** | **Total** | **Comment** |
| --- | --- | --- |
| Staff costs | £84,377 |  |
| Room hire | £26,857 | =400 (room hire cost)*67.14 (number of days); assumes cost of hire is £400 / day |
| Consumables | £15,839 | Consumables include costs associated with printing, paper, classroom |
| Overheads | £37,970 | Direct overheads based on 29% of direct care salary costs and indirect overheads based on 16% direct care salary costs (As used in Curtis et al 2019 for overheads for healthcare professionals) |
| Maintenance of app | £16,200 | See above table |
| Capital cost, development of app | £3240 | See above |
| Capital cost, development of EACH-B | £8745 | See above |

**Table 11: Total costs**

| **Item** | **Total** | **Comment** |
| --- | --- | --- |
| Total cost | £193,228 | Sum of all of the above costs |
| Cost per pupil | £154.58 | =£193,228/1250; (total cost /total number of pupils) |

3 Appendix C

**Figure 1: Cost-Effectiveness Acceptability Curve (CEAC)**

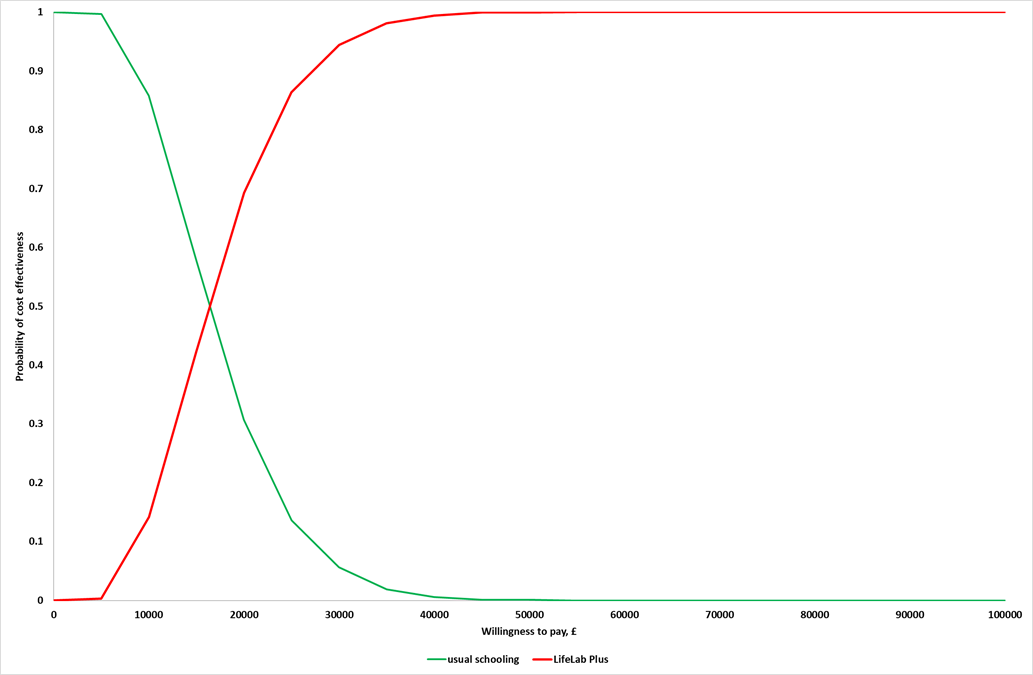
